# Association between maternal body mass index, skin incision-to-delivery time, and umbilical artery pH in cesarean deliveries

**DOI:** 10.1101/2025.04.20.25326121

**Authors:** Nadine Sunji, Alyssa M. Hernandez, Rachel Schmidt, Amy Y. Pan, Nina Ayala, Margaret H. Bublitz, Anna Palatnik

**Affiliations:** Department of Obstetrics and Gynecology, Medical College of Wisconsin, Milwaukee, Wisconsin, United States; Department of Pediatrics, Medical College of Wisconsin, Milwaukee, Wisconsin, United States; Department of Obstetrics and Gynecology, Alpert Medical School of Brown University, Providence, Rhode Island, United States; Department of Psychiatry and Human Behavior, Alpert Medical School of Brown University, Providence, Rhode Island, United States; Department of Medicine, Alpert Medical School of Brown University, Providence, Rhode Island, United States; Behavioral and Social Sciences, Brown School of Public Health, Providence, Rhode Island, United States; Cardiovascular Center, Medical College of Wisconsin, Milwaukee, Wisconsin, United States

**Keywords:** cesarean delivery, neonatal acidosis, obesity, umbilical artery, body mass index, operative time

## Abstract

**Objective:** To estimate the association between maternal body mass index (BMI) at delivery, time from skin incision to infant delivery, and umbilical artery (UA) pH <7.0.

**Methods:** This was a secondary analysis of the Assessment of Perinatal Excellence, a multicenter observational study of an obstetrical cohort of individuals who delivered between 2008 and 2011 in the United States. This analysis included women who delivered via cesarean with known BMI at delivery, skin incision-to-delivery time, and UA pH. Multivariable linear regression assessed the association between BMI and time from skin incision to infant delivery while multivariable logistic regression estimated the associations of BMI and time from skin incision to delivery with UA pH <7.0. An interaction between BMI and skin incision-to-delivery time was evaluated to examine their combined effect on UA pH <7.0.

**Results:** A total of 16,723 women were included across five BMI groups. Increasing BMI was associated with longer time intervals from skin incision to delivery and higher rates of UA pH <7.0. After controlling for potentially confounding factors, all BMI groups ≥25 kg/m^2^ were associated with longer time intervals from skin incision to delivery. Specifically, BMI groups of 40-49.9 kg/m^2^ and ≥50 kg/m^2^ had skin incision-to-delivery times that were 1.30 (95% CI 1.23-1.36) and 1.44 (95% CI 1.34-1.55) times longer, respectively, compared with BMI <25 kg/m^2^. In the multivariable logistic regression, BMI group ≥50 kg/m^2^ remained associated with higher odds of UA pH <7.0. There was a significant interaction between BMI and time from skin incision to delivery regarding the risk of UA pH <7.0 (p for the interaction term=.003).

**Conclusion:** Maternal BMI ≥50 kg/m^2^ was associated with longer time from skin incision to infant delivery and higher odds of UA pH <7.0. Skin incision-to-delivery time had a differential impact on UA pH in each BMI group.

**Precis:** In cesarean deliveries, maternal body mass index ≥50 kg/m^2^ and prolonged skin incision-to-delivery time increase the risk of umbilical artery pH <7.0.

## Introduction

Rates of obesity continue to rise in the United States, especially in the obstetric population. Between 2016 and 2019, pre-pregnancy obesity, defined as body mass index (BMI) ≥30 kg/m^2^, increased from 26.1% to 29.0% among women of reproductive age.^1^ Obesity in pregnancy increases the risk of multiple adverse maternal and neonatal outcomes, including pregnancy loss, gestational diabetes, hypertensive disorders of pregnancy, macrosomia, stillbirth, and congenital anomalies.^2^ Parturients with obesity also face a higher risk of labor dystocia and cesarean delivery.^3,4^ Cesarean delivery in this patient population carries a higher risk of surgical complications, such as neuraxial anesthesia failure, excessive blood loss, wound dehiscence, and surgical site infection.^5^

Studies have demonstrated prolonged operative times in pregnant women with obesity,^6,7^ a few of which report a significant association between increasing BMI and longer skin incision-to-delivery times.^8,9^ This delay may be due to mechanical challenges, including difficulty achieving adequate surgical exposure, increased vascularity causing excessive bleeding, and inadequate anesthesia.^10^ While prolonged skin incision-to-delivery times have been observed, the impact on neonatal outcomes remains unclear. Skin incision-to-delivery time may represent a critical period that directly affects fetal oxygenation and acid-base balance.^8,9^ Neonatal acidosis, a marker of fetal hypoxia, has been associated with an increased risk of adverse outcomes, including neonatal intensive care unit (NICU) admission, hypoxic-ischemic encephalopathy, and neurodevelopmental delays.^11,12^ However, the extent to which maternal BMI and prolonged skin incision-to-delivery time contribute to the risk of neonatal acidosis has not been well characterized. Therefore, this study aimed to assess the relationship between maternal BMI and skin incision-to-delivery time as well as their combined effect on the risk of neonatal acidosis.

## Methods

This was a secondary analysis of the Assessment of Perinatal Excellence (APEX) cohort consisting of 115,502 individuals who delivered live infants ≥23 weeks’ gestation at hospitals participating in the Eunice Kennedy Shriver National Institute of Child Health and Human Development (NICHD) Maternal-Fetal Medicine Units (MFMU) Network between March 2008 and February 2011. To explore potential quality measures of obstetric care in the intrapartum period, the APEX study used chart abstraction to collect data on demographic, social, medical, and obstetric history, as well as pregnancy and neonatal outcomes. Details of the APEX study design and methodology have been reported in a previous publication.^13^ This secondary analysis was approved by the Institutional Review Board at the Medical College of Wisconsin and the Eunice Kennedy Shriver NICHD.

Our study included women who delivered via cesarean with available data on height and weight at delivery, time of skin incision, time of infant delivery, and umbilical artery (UA) pH. The analysis excluded those with combined vaginal-cesarean delivery, intrauterine fetal demise, placenta accreta, or uterine rupture as these conditions substantially affect study outcomes. The parturient’s height and most recent pregnancy weight were used to calculate BMI. Women who met inclusion criteria were then stratified into five groups defined by BMI at delivery: <25, 25-29.9, 30-39.9, 40-49.9, and ≥50 kg/m^2^. BMI <25 kg/m^2^ served as the referent group.

The primary outcome of this study was time from skin incision to infant delivery, which was derived by subtracting the time of skin incision from the birth time. The secondary outcome was UA pH <7.0. Maternal sociodemographic characteristics, delivery characteristics, and pregnancy outcomes were extracted from the APEX dataset and compared among the five BMI groups.

Chi-square test and Fisher’s exact test were used to compare categorical variables, and Kruskal-Wallis test was used to compare continuous variables. The Cochran-Armitage trend test was used to examine the linear trend in binomial proportions across BMI categories while Spearman correlation coefficient was used to determine the relationship between continuous variables and BMI categories. A general linear model was performed to assess the association between time from skin incision to infant delivery and BMI categories. Time from skin incision to infant delivery was log transformed and then back transformed to the raw scale, and the geometric mean ratio and 95% CI were calculated between each BMI group and the referent group. A logistic regression was used to estimate the associations of time from skin incision to delivery and BMI categories with UA pH <7.0. An interaction term of time from skin incision to delivery and BMI categories was also included to examine their combined effect on the outcome of UA pH <7.0. Potential confounders, chosen based on our univariate analyses, clinical expertise, and variables identified in prior studies to be predictors of the time interval from skin incision to infant delivery, were included in the multivariable analysis. All tests were two-tailed, and a p-value below 0.05 defined statistical significance. Dunnett’s test and the False Discovery Rate (FDR) method were used to adjust for multiple comparisons. All analyses were performed with the software SAS version 9.4 (SAS Institute Inc., Cary, NC) and StatXact-12.

## Results

Of the 115,502 women in the original APEX cohort, 16,723 met inclusion criteria and were included across five BMI groups in this secondary analysis. When stratified by BMI at delivery, 1,466 (8.8%) patients had a BMI <25 kg/m^2^, 4,828 (28.9%) had a BMI 25-29.9 kg/m^2^, 7,836 (46.9%) had a BMI 30-39.9 kg/m^2^, 2,061 (12.3%) had a BMI 40-49.9 kg/m^2^, and 532 (3.2%) had a BMI ≥50 kg/m^2^. Table 1 summarizes baseline maternal characteristics stratified by BMI group. Patients significantly differed across all characteristics except alcohol use during pregnancy. Increasing BMI was associated with Black or African American race, multiparity, government insurance, tobacco use during pregnancy, singleton gestation (p=.002), prior cesarean delivery, hypertensive disorders of pregnancy, and diabetes (all otherwise p<.001).

**Table 1.** Baseline maternal characteristics stratified by body mass index at delivery.

| Characteristic | BMI <25<br>(n = 1466) | BMI 25-29.9<br>(n = 4828) | BMI 30-39.9<br>(n = 7836) | BMI 40-49.9<br>(n = 2061) | BMI ≥50<br>(n = 532) | p-value |
| --- | --- | --- | --- | --- | --- | --- |
| Maternal age (years) |  |  |  |  |  | <.001 |
| <20 | 163 (11.1) | 371 (7.7) | 507 (6.5) | 129 (6.3) | 32 (6.0) |  |
| 20-24 | 328 (22.4) | 818 (16.9) | 1488 (19.0) | 476 (23.1) | 115 (21.6) |  |
| 25-29 | 312 (21.3) | 1070 (22.2) | 2022 (25.8) | 585 (28.4) | 147 (27.6) |  |
| 30-34 | 361 (24.6) | 1370 (28.4) | 2083 (26.6) | 479 (23.2) | 138 (25.9) |  |
| ≥35 | 302 (20.6) | 1199 (24.8) | 1736 (22.2) | 392 (19.0) | 100 (18.8) |  |
| Race |  |  |  |  |  | <.001 |
| American<br>Indian/Alaska<br>Native | 2 (0.1) | 6 (0.1) | 13 (0.2) | 9 (0.4) | 2 (0.4) |  |
| Asian | 88 (6.0) | 303 (6.3) | 187 (2.4) | 19 (0.9) | 6 (1.1) |  |
| Black/African<br>American | 318 (21.7) | 872 (18.1) | 1939 (24.7) | 809 (39.3) | 291 (54.7) |  |
| Native<br>Hawaiian/Other<br>Pacific Islander | 2 (0.1) | 11 (0.2) | 14 (0.2) | 9 (0.4) | 2 (0.4) |  |
| White | 828 (56.5) | 2692 (55.8) | 3981 (50.8) | 888 (43.1) | 141 (26.5) |  |
| None of the<br>above | 176 (12.0) | 699 (14.5) | 1333 (17.0) | 265 (12.9) | 78 (14.7) |  |
| Not documented | 52 (3.6) | 245 (5.1) | 369 (4.7) | 62 (3.0) | 12 (2.3) |  |
| Hispanic or Latino<br>ethnicity | 294 (20.1) | 1241 (25.7) | 2560 (32.7) | 516 (25.0) | 96 (18.1) | <.001 |
| Nulliparous | 659 (45.0) | 2267 (47.0) | 3249 (41.5) | 792 (38.5) | 208 (39.1) | <.001 |
| Health insurance |  |  |  |  |  | <.001 |
| Private | 674 (46.3) | 2436 (51.1) | 3261 (42.0) | 699 (34.2) | 144 (27.2) |  |
| Government | 642 (44.1) | 1760 (37.0) | 3499 (45.1) | 1152 (56.3) | 352 (66.4) |  |
| Uninsured or<br>self-pay | 141 (9.7) | 568 (11.9) | 1004 (12.9) | 195 (9.5) | 34 (6.4) |  |
| Tobacco use during<br>pregnancy | 187 (12.8) | 407 (8.4) | 748 (9.6) | 279 (13.6) | 90 (16.9) | <.001 |
| Alcohol use during<br>pregnancy | 43 (2.9) | 132 (2.7) | 222 (2.8) | 55 (2.7) | 20 (3.8) | .723 |
| Illicit drug use<br>during pregnancy | 95 (6.5) | 199 (4.1) | 311 (4.0) | 94 (4.6) | 35 (6.6) | <.001 |
| Multifetal gestation | 168 (11.5) | 540 (11.2) | 766 (9.8) | 185 (9.0) | 53 (10.0) | .014 |
| Prior cesarean<br>deliveries | 482 (49.0) | 1708 (53.6) | 3202 (57.9) | 861 (57.8) | 226 (58.9) | <.001 |
| Hypertensive<br>disorders of<br>pregnancy | 174 (11.9) | 601 (12.5) | 1614 (20.6) | 717 (34.8) | 265 (49.8) | <.001 |
| Diabetes |  |  |  |  |  | <.001 |
| Pregestational | 12 (0.8) | 80 (1.7) | 279 (3.6) | 149 (7.2) | 61 (11.5) |  |
| Gestational | 54 (3.7) | 218 (4.5) | 704 (9.0) | 269 (13.1) | 87 (16.4) |  |

Abbreviations: BMI, body mass index.
Data shown as n (%).

All delivery outcomes also significantly differed between BMI groups (Table 2). Total operative time increased from a median of 45 (interquartile range [IQR] 35-57) minutes in the BMI <25 and 25-29.9 kg/m^2^ groups to 48 (IQR 38-61) minutes in the BMI 30-39.9 kg/m^2^ group, 54 (IQR 42-68) minutes in the BMI 40-49.9 kg/m^2^ group, and 62 (IQR 48-79) minutes in the BMI ≥50 kg/m^2^ group (p<.001). Rates of unscheduled and emergency cesarean deliveries were more likely to be lower in higher BMI groups (p<.001). The indication for cesarean delivery was more likely to be non-reassuring fetal status in lower BMI groups but labor dystocia in higher BMI groups (p<.001). Vertical skin incisions and classical uterine incisions were more likely in higher BMI groups (p<.001). Infant birth weight was more likely to increase as BMI increased (p<.001). Infants of those with a BMI <25 kg/m^2^ had the lowest gestational age at delivery compared with all other groups (p<.001) and the highest rates of Apgar scores <7 at 5 minutes and NICU/intermediate nursery admissions (p<.001 for both outcomes).

**Table 2.** Delivery outcomes stratified by body mass index at delivery.

| **Delivery outcome** | **BMI <25**<br>**(n = 1466)** | **BMI 25-**<br>**29.9**<br>**(n = 4828)** | **BMI 30-**<br>**39.9**<br>**(n = 7836)** | **BMI 40-**<br>**49.9**<br>**(n = 2061)** | **BMI ≥50**<br>**(n = 532)** | **p-**<br>**value** |
| --- | --- | --- | --- | --- | --- | --- |
| Time from skin incision to infant delivery (minutes) | 8 (5-11) | 8 (5-12) | 9 (6-14) | 11 (7-16) | 12 (8-18) | <.001 |
| Total operative time (minutes) | 45 (35-57) | 45 (35-57) | 48 (38-61) | 54 (42-68) | 62 (48-79) | <.001 |
| Unscheduled cesarean | 1104 (75.3) | 3403 (70.5) | 5441 (69.4) | 1454 (70.6) | 343 (64.5) | <.001 |
| Emergency cesarean | 318 (28.8) | 880 (25.9) | 1116 (20.5) | 269 (18.5) | 66 (19.2) | <.001 |
| Indication for current cesarean |  |  |  |  |  | <.001 |
| Non-reassuring fetal status | 343 (23.4) | 1104 (22.9) | 1646 (21.0) | 440 (21.4) | 111 (20.9) |  |
| Labor dystocia (including failed induction) | 202 (13.8) | 955 (19.8) | 1670 (21.3) | 446 (21.7) | 105 (19.8) |  |
| Type of skin incision |  |  |  |  |  | <.001 |
| Pfannenstiel | 1299 (89.3) | 4262 (89.0) | 6656 (85.7) | 1763 (86.3) | 418 (78.9) |  |
| Vertical | 156 (10.7) | 525 (11.0) | 1108 (14.3) | 280 (13.7) | 112 (21.1) |  |
| Type of uterine incision |  |  |  |  |  | <.001 |
| Low transverse | 1360 (93.3) | 4587 (95.3) | 7432 (95.3) | 1915 (93.2) | 455 (86.2) |  |
| Classical, other vertical | 64 (4.4) | 123 (2.6) | 196 (2.5) | 75 (3.7) | 55 (10.4) |  |
| Low vertical | 24 (1.7) | 88 (1.8) | 130 (1.7) | 45 (2.2) | 11 (2.1) |  |
| T or J incision | 9 (0.6) | 15 (0.3) | 42 (0.5) | 19 (0.9) | 7 (1.3) |  |
| Type of anesthesia |  |  |  |  |  | <.001 |
| Epidural | 558 (38.1) | 2147 (44.5) | 3541 (45.2) | 972 (47.2) | 269 (50.7) |  |
| Spinal | 1002 (68.4) | 3229 (67.0) | 5231 (66.8) | 1298 (63.0) | 314 (59.1) |  |
| General | 144 (9.8) | 362 (7.5) | 474 (6.1) | 149 (7.2) | 43 (8.1) |  |
| Gestational age at delivery (weeks) | 38.0<br>(34.6-39.3) | 39.0<br>(37.0-39.9) | 39.0<br>(37.1-39.7) | 38.7<br>(37.0-39.6) | 38.6<br>(36.6-39.4) | <.001 |
| Birth weight (grams) | 2793<br>(2080 -3235) | 3140<br>(2615-3515) | 3260<br>(2780-3665) | 3275<br>(2745-3737) | 3250<br>(2689-3746) | <.001 |
| 5-minute Apgar score <7 | 86 (5.9) | 178 (3.7) | 265 (3.4) | 82 (4.0) | 24 (4.5) | <.001 |
| Umbilical artery pH <7.0 | 14 (1.0) | 76 (1.6) | 120 (1.5) | 46 (2.2) | 21 (4.0) | <.001 |
| NICU/intermediate nursery admission | 582 (39.7) | 1329 (27.5) | 2023 (25.8) | 581 (28.2) | 160 (30.1) | <.001 |

Abbreviations: BMI, body mass index; NICU, neonatal intensive care unit.
Data shown as n (%) or median (interquartile range).

Both time from skin incision to infant delivery and rates of UA pH <7.0 significantly increased as BMI increased (Table 2). The median time from skin incision to infant delivery was 8 (IQR 5-11) minutes in the BMI <25 kg/m^2^ group, 8 (IQR 5-12) minutes in the BMI 25-29.9 kg/m^2^ group, 9 (IQR 6-14) minutes in the BMI 30-39.9 kg/m^2^ group, 11 (IQR 7-16) minutes in the BMI 40-49.9 kg/m^2^ group, and 12 (IQR 8-18) minutes in the BMI ≥50 kg/m^2^ group (p<.001). Rates of UA pH <7.0 significantly increased from 1.0% in the BMI <25 kg/m^2^ group to 1.6% in the BMI 25-29.9 kg/m^2^ group, 1.5% in the BMI 30-39.9 kg/m^2^ group, 2.2% in the BMI 40-49.9 kg/m^2^ group, and 4.0% in the BMI ≥50 kg/m^2^ group (p<.001).

In the multivariable linear regression adjusting for maternal age, nulliparity, prior cesarean delivery, type of skin incision, type of uterine incision, scheduled versus unscheduled cesarean delivery, general anesthesia, hypertensive disorders of pregnancy, and diabetes, all BMI groups ≥25 kg/m^2^ remained significantly associated with longer times from skin incision to delivery (Table 3). Specifically, BMI groups of 25-29.9, 30-39.9, 40-49.9, and ≥50 kg/m^2^ had skin incision-to-delivery times that were 1.05 (95% CI 1.01-1.10), 1.14 (95% CI 1.10-1.19), 1.30 (95% CI 1.23-1.36), and 1.44 (95% CI 1.34-1.55) times longer, respectively, compared with BMI group <25 kg/m^2^.

**Table 3.** Association between maternal body mass index, skin incision-to-delivery time, and umbilical artery pH <7.0.

| **GMRs for skin incision-to-delivery time** |  |  |  |
| --- | --- | --- | --- |
| **BMI category** | **Unadjusted GMR (95% CI)** | **Adjusted GMR\* (95% CI)** | **Dunnett adjusted p-value** |
| BMI <25 | Referent | Referent | -- |
| BMI 25-29.9 | 1.08 (1.04-1.12) | 1.05 (1.01-1.10) | .049 |
| BMI 30-39.9 | 1.21 (1.16-1.25) | 1.14 (1.10-1.19) | <.001 |
| BMI 40-49.9 | 1.38 (1.32-1.44) | 1.30 (1.23-1.36) | <.001 |
| BMI ≥50 | 1.59 (1.49-1.69) | 1.44 (1.34-1.55) | <.001 |
| **ORs for umbilical artery pH <7.0** |  |  |  |
| **Variable** | **Unadjusted OR (95% CI)** | **Adjusted OR\* (95% CI)** | **p-value†** |
| BMI |  |  | .025 |
| BMI <25 | Referent | Referent | -- |
| BMI 25-29.9 | 1.71 (0.96-3.03) | 1.69 (0.85-3.38) | .136 |
| BMI 30-39.9 | 1.76 (1.01-3.06) | 1.73 (0.89-3.39) | .136 |
| BMI 40-49.9 | 2.76 (1.51-5.06) | 2.26 (1.09-4.70) | .057 |
| BMI ≥50 | 5.39 (2.70-10.75) | 3.73 (1.60-8.70) | .010 |
| Skin incision-to-delivery time | 0.95 (0.93-0.97) | 1.00 (0.98-1.02) | .929 |

Abbreviations: GMR, geometric mean ratio; BMI, body mass index; OR, odds ratio; CI, confidence interval.
\* Adjusted for maternal age, nulliparity, prior cesarean, type of skin incision, type of uterine incision, scheduled versus unscheduled cesarean delivery, general anesthesia, hypertensive disorders of pregnancy, and diabetes.
† P-values for BMI categories were adjusted post hoc for multiple comparisons using the False Discovery Rate method.

In the multivariable logistic regression (Table 3), BMI group ≥50 kg/m^2^ remained associated with higher odds of UA pH <7.0 after controlling for multiple comparisons with the FDR method (adjusted odds ratio [aOR] = 3.73, 95% CI 1.60-8.70). Skin incision-to-delivery time was not associated with UA pH <7.0 (aOR = 1.00, 95% CI 0.98-1.02). However, an interaction between BMI and skin incision-to-delivery time was noted and remained statistically significant after controlling for confounders (interaction p=.003). Table 4 shows the aORs and 95% CIs for various BMI categories at the study population’s mean skin incision-to-delivery interval (10.3 minutes), median (9.0 minutes), lower quartile (6.0 minutes), and upper quartile (13.0 minutes). BMI groups 40-49.9 and ≥50 kg/m^2^ remained significantly associated with higher odds of UA pH <7.0 at all skin incision-to-delivery time intervals, and BMI group 30-39.9 kg/m^2^ was significantly associated with higher odds of UA pH <7.0 at skin incision-to-delivery time intervals of 9 (aOR = 4.51, 95% CI 1.15-17.74), 10.3 (aOR = 6.47, 95% CI 1.28-32.61), and 13 minutes (aOR = 13.68, 95% CI 1.58-118.64).

**Table 4.** Odds of umbilical artery pH <7.0 by maternal body mass index and skin incision-to-delivery time.

| BMI category | 6.0 minutes |  | 9.0 minutes |  | 10.3 minutes |  | 13.0 minutes |  |
| --- | --- | --- | --- | --- | --- | --- | --- | --- |
|  | OR (95% CI) | aOR* (95% CI) | OR (95% CI) | aOR* (95% CI) | OR (95% CI) | aOR* (95% CI) | OR (95% CI) | aOR* (95% CI) |
| **BMI <25** | Referent | Referent | Referent | Referent | Referent | Referent | Referent | Referent |
| **BMI 25-29.9** | 2.15<br>(1.09-4.24) | 2.33<br>(0.96-5.65) | 3.05<br>(1.05-8.83) | 3.96<br>(0.99-15.90) | 3.55<br>(1.00-12.58) | 4.98<br>(0.96-25.84) | 4.86<br>(0.88-26.96) | 8.03<br>(0.89-72.90) |
| **BMI 30-39.9** | 1.89<br>(0.97-3.71) | 1.96<br>(0.81-4.73) | 4.09<br>(1.45-11.53) | 4.51<br>(1.15-17.74) | 5.71<br>(1.67-19.53) | 6.47<br>(1.28-32.61) | 11.43<br>(2.18-59.86) | 13.68<br>(1.58-118.64) |
| **BMI 40-49.9** | 3.17<br>(1.53-6.57) | 3.00<br>(1.17-7.70) | 6.43<br>(2.22-18.59) | 6.28<br>(1.55-25.49) | 8.73<br>(2.50-30.47) | 8.65<br>(1.67-44.73) | 16.49<br>(3.09-87.95) | 16.82<br>(1.90-148.86) |
| **BMI ≥50** | 3.99<br>(1.65-9.61) | 3.30<br>(1.07-10.12) | 9.69<br>(3.08-30.43) | 7.99<br>(1.78-35.94) | 14.23<br>(3.84-52.77) | 11.72<br>(2.09-65.64) | 31.65<br>(5.76-173.73) | 25.99<br>(2.80-241.06) |

Abbreviations: BMI, body mass index; OR, odds ratio; CI, confidence interval; aOR, adjusted odds ratio.
\* Adjusted for maternal age, nulliparity, prior cesarean, type of skin incision, type of uterine incision, scheduled versus unscheduled cesarean delivery, general anesthesia, hypertensive disorders of pregnancy, and diabetes.

## Discussion

In this secondary analysis of the APEX study, we investigated the association between maternal BMI at delivery, time from skin incision to infant delivery, and UA pH <7.0. We found that increasing maternal BMI was associated with increasing time from skin incision to delivery in all BMI groups and with UA pH <7.0 in BMI group ≥50 kg/m^2^. The association between skin incision-to-delivery time and UA pH <7.0 did not persist after adjusting for confounding variables. However, a significant interaction between BMI and skin incision-to-delivery time towards UA pH <7.0 was detected, suggesting that the combination of higher BMI and longer skin incision-to-delivery time increases the risk of UA pH <7.0 more than higher BMI alone.

Consistent with our findings, previous studies have reported that time from skin incision to delivery significantly increases with increasing BMI.^9,14–17^ In a large cohort from the MFMU Cesarean Registry, Girsen *et al.*^9^ demonstrated that BMI ≥30 kg/m^2^ significantly increased the risk of a skin incision-to-delivery time of ≥18 minutes by about 2-3 times compared with BMI 18.5-24.9 kg/m^2^ after adjusting for confounders. Similarly, Zewdu *et al.*^17^ found a 5.38-fold increase in the odds of a prolonged skin incision-to-delivery time >8 minutes in individuals with a BMI >30 kg/m^2^. Another study using data from a single U.S. institution showed that time from skin incision to delivery significantly increased linearly with increasing BMI.^8^

Our study also identified a significant association between BMI ≥50 and UA pH <7.0, consistent with previous studies reporting an inverse relationship between BMI and UA pH.^8,9,14–16,18–22^ Edwards *et al.*^20^ reported a 0.1-decrease in UA pH with each 10-unit increase in BMI in low-risk, nonlaboring individuals who delivered via cesarean at term and received spinal anesthesia. Most studies investigated higher pH thresholds (<7.1 or <7.2) likely due to smaller sample sizes that further limited the ability to evaluate more severe acidosis. For example, Ituk *et al.*^23^ did not identify a significant association between increasing BMI and UA pH <7.1, possibly due to the low frequency of this outcome in their sample. We specifically examined UA pH <7.0 as it is more strongly associated with neonatal morbidity.^11,24^

Like previous smaller studies,^25,26^ we did not detect an association between skin incision- to-delivery time and UA pH <7.0. Different indications for a cesarean delivery may influence acid-base balance, rather than skin incision-to-delivery time alone, as maternal and fetal conditions would differ at baseline. In elective or non-emergent cesarean deliveries, the fetus is typically stable at baseline and can therefore tolerate longer operative time intervals.^26^ Fetal distress at baseline, however, may predispose to lower UA pH regardless of any intraoperative time interval.^27^ Stable maternal hemodynamics may also better maintain fetal perfusion and oxygenation and mitigate the impact of prolonged surgical time intervals. In scheduled term cesarean deliveries, Knigin *et al.*^14^ demonstrated a nearly threefold increased risk of umbilical arterial pH ≤7.1 when skin incision-to-delivery time exceeded 13 minutes and a 1.7-fold increased risk at >7 minutes. Our study, in contrast, examined severe neonatal acidosis, defined as UA pH <7.0, and controlled for scheduled versus unscheduled cesarean deliveries rather than restricting our sample to either delivery type. Because Knigin *et al.*^14^ examined a higher pH threshold, they may have detected a subtler effect of skin incision-to-delivery time on UA pH, particularly in planned cesarean deliveries where the likelihood of adverse neonatal outcomes is lower compared with unscheduled cesarean deliveries.

A key finding in our study was the significant interaction between maternal BMI and skin incision-to-delivery time, demonstrating that skin incision-to-delivery time had a varying impact on UA pH in each BMI group. We found that the odds of UA pH <7.0 increased with both higher BMI and prolonged skin incision-to-delivery time. For instance, in individuals with BMI ≥50 kg/m^2^, the odds of UA pH <7.0 were 3.30 times higher than in those with BMI <25 kg/m^2^ when skin incision-to-delivery time was 6 minutes. When skin incision-to-delivery time was 13 minutes, the odds increased to 25.99. This suggests that the impact of BMI on UA pH varies at different time intervals, reinforcing the need for more efficient surgical approaches in individuals with higher BMI in addition to obesity risk reduction and management.

Although not directly explored in our study, prior research suggests that intraoperative hemodynamics may contribute to the relationship between BMI and UA pH.^15,21^ In scheduled cesarean deliveries with spinal anesthesia, Powell *et al.*^15^ found that the inverse association between BMI and UA pH did not persist after adjusting for confounders, including intraoperative blood pressure, suggesting that intraoperative hypotension may explain the relationship between increasing BMI and decreasing UA pH. However, spinal anesthesia is more commonly associated with neonatal acidosis than general or epidural anesthesia,^28^ and our study included but controlled for all types of anesthesia as a potential confounder, making our findings more broadly applicable to the general obstetric population.

Based on our findings, minimizing skin incision-to-delivery time may largely mitigate the risk of severe neonatal acidosis particularly in individuals with BMI ≥50 kg/m^2^. Time from skin incision to delivery did not appear to affect the risk of UA pH <7.0 in those with BMI <30 kg/m^2^. Therefore, a tailored protocol for surgical planning of cesarean deliveries in patients with BMI ≥50 kg/m^2^ that may improve access to the incision site or hysterotomy could be developed and optimized to reduce skin incision-to-delivery time. Preoperative patient counseling should include careful inspection of the abdominal wall and a discussion with the patient regarding the type of skin incision given that over 20% of patients with BMI ≥50 kg/m^2^ in this cohort had a vertical skin incision. Our results could also factor into individualized risk assessment for fetal acidemia during cesarean delivery and support intrapartum clinical decision-making, potentially prompting a lower threshold to move towards cesarean delivery in the setting of prolonged category II fetal heart rate tracings in patients with BMI ≥50 kg/m^2^. Lastly, our findings underscore the importance of obesity prevention and management through prenatal counseling, nutrition, behavioral support, and physical activity. Clinicians should inform patients with obesity about delivery options and associated risks.

With the substantial increase in the prevalence of obesity among individuals of reproductive age, further research should focus on identifying surgical techniques that may reduce the skin incision-to-delivery time in patients with higher BMIs. Obstetric interventions, such as early induction of labor, can also be investigated to assess their impact on neonatal outcomes in this population. In a Canadian cohort with BMI ≥30 kg/m^2^, planned cesarean delivery was associated with a 30% reduction in the risk of adverse neonatal outcomes compared with spontaneous labor, but there was no significant difference when comparing labor induction with spontaneous labor.^29^ To determine the optimal mode of delivery in pregnancies complicated by obesity, further studies can compare neonatal outcomes in scheduled versus emergent cesarean deliveries in a U.S. population. This may elucidate when the benefits of a planned cesarean delivery outweigh the risks of an emergent delivery.

There are several strengths to this study. To our knowledge, this is the largest study evaluating the effect of both maternal BMI and skin incision-to-delivery time on UA pH. The APEX study’s large cohort size and access to UA pH data strengthen its validity, addressing a gap in the literature by providing robust, generalizable evidence across BMI categories.

Furthermore, our large sample consisted of individuals from multiple medical centers throughout the U.S., thus increasing generalizability, and women with BMI ≥30 kg/m^2^ comprised more than half of our sample. We were also able to adjust for many possibly confounding factors, including incision type, indication for cesarean delivery, and comorbidities frequently present in higher BMI groups, so our results may be more clinically relevant and generalizable.

This study also has some limitations. We restricted the original APEX sample to those with available data on UA pH, which decreased the sample size available for analysis and may have introduced selection bias. Based on the APEX study design, standard protocols were not altered across the U.S. hospitals; therefore, UA gases were obtained based on institutional or individual provider practice. Nevertheless, the remaining sample of 16,723 individuals was still robust with 277 (1.7%) of patients exhibiting UA pH <7.0, which is slightly higher than previously reported incidence rates ranging from 0.26% to 1.3% but remains within a comparable range.^30^ Stratifying cases with UA pH <7.0 into five BMI groups resulted in smaller sample sizes, which may have led to wider CIs and less precise estimates in our interaction analysis. Finally, while we detected an association between maternal BMI ≥50 kg/m^2^ and UA pH <7.0, we did not account for intraoperative hypotension, which has been suggested as a possible explanation for the association between increasing BMI and decreasing UA pH.^15,21^

In conclusion, our findings demonstrate that skin incision-to-delivery time increases with increasing BMI and that BMI ≥50 kg/m^2^ increased the risk of UA pH <7.0 in cesarean deliveries. We also found that BMI ≥50 kg/m^2^ and prolonged skin incision-to-delivery time jointly increase the risk of UA pH <7.0. These results highlight the importance of individualized risk assessment and comprehensive prenatal obesity management including tailored surgical planning to optimize maternal and neonatal outcomes.

## Data Availability

All data produced are available online at DASH NICHD website. This was a secondary analysis of that data set.

https://dash.nichd.nih.gov/study/424640

The authors declare that they have no known competing financial interests or personal relationships that could have appeared to influence the work reported in this paper.

We are grateful to the study investigators and research staff who were involved with An Observational Cohort Study to Evaluate Measures of Quality of Obstetric Care - Assessment of Perinatal Excellence (APEX). In particular, we acknowledge the assistance of the Maternal-Fetal Medicine Units Network, the Eunice Kennedy Shriver National Institute of Child Health and Human Development, and the APEX Protocol Subcommittee in making the database available. The contents of this report represent the views of the authors and do not represent the views of the Eunice Kennedy Shriver National Institute of Child Health and Human Development or the National Institutes of Health.

We acknowledge NICHD DASH for providing the APEX Registry: Assessment of Perinatal Excellence data that was used for this research.

Findings of this study were presented at the 72^nd^ Annual Meeting of the Society for Reproductive Investigation, Charlotte, North Carolina, United States, March 25-29, 2025.

